# Multiverse to inform neurological research: an example using recovery outcome of neglect

**DOI:** 10.1101/2021.03.29.21254222

**Authors:** Margaret J Moore, Nele Demeyere

**Author notes:** ^*^**Corresponding Author:** Prof. Nele Demeyere. **Author Contributions:** MJM conducted analysis and drafted the manuscript. ND conceptualised and supervised the study, acquired funding and edited manuscript drafts.

## Abstract

**Objective:** Multiverse analysis provides an ideal tool for understanding how inherent, yet ultimately arbitrary methodological choices impact the conclusions of individual studies. With this investigation we aimed to demonstrate the utility of multiverse analysis for evaluating generalisability and identifying potential sources of bias within studies employing neurological populations.

**Method:** Multiverse analysis was used to evaluate the robustness of the relationship between post-stroke visuospatial neglect and poor long term recovery outcome within a sample of 1113 (age =72.5, 45.1% female) stroke survivors. A total of 25,600 t-test comparisons were run across 400 different patient groups defined using various combinations of valid inclusion criteria based on lesion location, stroke type, assessment time, neglect impairment definition, and scoring criteria across 16 standardised outcome measures.

**Results:** Overall, 33.9% of conducted comparisons yielded significant results. 99.9% of these significant results fell below the null specification curve, indicating a highly robust relationship between neglect and poor recovery outcome. However, the strength of this effect was not constant across all comparison groups. Comparisons which included <100 participants, pre-selected patients based on lesion type, or failed to account for allocentric neglect impairment were found to yield average effect sizes which differed substantially. Similarly, average effect sizes differed across various outcome measures with the strongest average effect in comparisons involving an activities of daily living measure and the weakest in comparisons employing a depression subscale.

**Conclusions:** This investigation demonstrates the utility of multiverse analysis techniques for evaluating effect robustness and identifying potential sources of bias within neurological research.

## Introduction

Conducting any research study inevitably involves choosing specific analysis designs, outcome measures, and variables of interest from a multitude of possible choices. It is often unclear how these inherent, yet arbitrary methodological choices ultimately impact the final conclusions of any single study. It is plausible that the results of any one individual analysis can be drastically skewed, and therefore unrepresentative due to the specific combination of methodological choices made by the researchers[1]. This possibility is critically important to consider within the context of any neurological research.

Planning research within neurological populations requires researchers to select specific combinations of potentially valid methodological choices. For example, studies aiming to investigate post-stroke impairment must choose between various standard inclusion criteria with some studies opting to include patients with specific stroke types and including a sample selected regardless of stroke characteristics. Similarly, neurological investigations are required to choose from a wide range of equally valid behavioural outcome measures. For example, dementia-related cognitive impairment can be quantified using any one of multiple cognitive screening tools including the Mini Mental State Examination, Montreal Cognitive Assessment, or the ACE-R[2–4]. Neurological studies can also differ in inclusion criteria around the precise time window post diagnosis. For example, clinical trials investigating the efficacy of Parkinson’s treatment have commonly chosen to exclusively recruit patients within the early or later stages of disease progression[5]. These necessary choices are an inherent component of designing research within neurological populations, though they can sometimes be arbitrary (e.g. definitions of acute, subacute and chronic stages in stroke vary between studies[6]. However, the potential impact of these choices is not well understood.

In recent years, required pre-registration of protocols and trial registers have provided some degree of protection from spurious conclusions being drawn due to non-hypothesis driven research[7]. However, these quality controls do no not prevent biases arising from employing patient selection criteria and specific methodological choices. Regardless of the topic being investigated, methodological choices can ultimately impact the final conclusions of any single study. In order to better understand potential biases in selection measures, secondary analyses should be informed by a clear understanding of potential biases in selection measures which can be gleaned from secondary analyses of existing large data sets can help inform choices for strong confirmatory research designs.

Multiverse analysis provides an ideal methodology for evaluating the impact of potentially arbitrary design choices on the conclusions of neurological research[1,8]. In a multiverse analysis, a large number of potential datasets are generated by making each possible permutation of reasonable methodological choices within a single experiment. Each of these datasets are then independently analysed, and the cumulative results are used to evaluate the overall robustness of the effect of interest. Critically, multiverse analysis clarifies which specific methodological choices may lead to significant effects. Multiverse analysis is not yet a common approach in research within neurological populations, even though this analysis technique helps in evaluating the potential impact of inclusion criteria, variables of interest, and key outcome measures. This increase in transparency can potentially help evaluate past research where documented effects may have been partly or wholly due to the specific outcome measures employed, or more clearly identify potential limits in generalising results to wider patient populations. Given this method’s potential for helping to evaluate the robustness of findings, more studies assessing the utility of multiverse analysis techniques within specific key questions addressed in neurological research are needed. Here, we provide an example multiverse analysis within post-stroke cognition research, specifically on the effect of acute visuo-spatial on longer term recovery outcomes, exploring the impact of decision making from our own study[9]

### Demonstration of Multiverse Analysis

Research investigating the predictive relationship between the occurrence of visuospatial neglect and poor long-term recovery outcome in stroke survivors provides an ideal context in which to investigate the utility of multiverse analysis within neurological research. Visuospatial neglect is a common cognitive deficit post-stroke, occurring in up to 48% of patients acutely[10], in which patients exhibit a marked inability to orient and attend to stimuli presented within one side of space[11]. A substantial body of evidence has found that the occurrence of acute visuospatial neglect acts as a strong predictor of poor recovery outcome in stroke survivor[12–14]. This association seems to be a particularly robust effect which is present within different neglect subtypes across multiple outcome measures, at multiple timepoints, and with varying inclusion criteria.

However, each investigation on this topic has made specific methodological choices which may potentially have impacted the study’s results. For example, some previous investigations of the relationship between acute visuospatial neglect and poor outcome have included a sample restricted to only patients with right hemisphere damage (e.g. [14]). While visuospatial neglect commonly occurs following both right and left hemisphere lesions, neglect following right hemisphere damage is typically more severe and long lasting [12,15-16]. Therefore, including only right hemisphere patients in visuospatial neglect studies could potentially result in a sample which is biased towards more severe neglect cases and it is not yet clear whether these restricted samples are adequately representative of neglect within the full stroke population.

Similarly, previous studies have included samples of neglect patients recruited at different timepoints following stroke (e.g.[12,17]). It has been demonstrated that neglect follows a non-linear recovery trajectory throughout the first six months following stroke, with most spontaneous recovery occurring within the first ten days following stroke[15]. This suggests that the “neglect” groups within more acute samples will include patients who will have improved by later timepoints and would therefore not be included in the “neglect” groups of studies who recruit patients at a later timepoint. It remains unclear whether and to what extent this difference in the specific patients included influenced the results of these studies.

Furthermore, visuospatial neglect is a highly heterogeneous condition with many different subtypes identified[18-19]. For example, some patients exhibiting impairment within an egocentric (self-centred) frame of reference and others within an allocentric (object-centred) reference frame [10, 20-21]. Egocentric and allocentric neglect represent doubly dissociated, independent cognitive impairments, though they also commonly co-occur within stroke survivors[10,22]. Many studies investigating the relationship between neglect and recovery outcome have exclusively included patients with egocentric neglect and have not included tests of allocentric neglect[13–15,17]. It therefore remains unclear whether studies which did not control for the potentially confounding impact of (additional or solely) allocentric neglect impairment produce externally valid conclusions.

Given that research investigating the predictive relationship between neglect and poor functional outcome in stroke survivors inherently requires such a wide range of methodological choices, this provides an ideal demonstration investigating the utility of multiverse analysis for evaluating the robustness and generalisability of effects within neurological research.

## Methods

This project represents a secondary analysis of longitudinal data collected as part of the Oxford Cognitive Screen (OCS) study (Demeyere et al 2015 - REC reference 11/WM/0299), OCS-Tablet Cognitive Screening study (REC reference 14/LO/0648) and OCS-Care study (Demeyere *et al*., 2019 – REC reference 12/WM/00335). All 3 studies included acute cognitive screening and 6 month follow ups. All patients provided informed consent in line with the Declaration of Helsinki.

### Participants

Participants were included in this investigation if they had completed the OCS Cancellation Task during initial hospitalisation and had completed at least one outcome measure during either acute hospitalisation or follow-up.

Importantly, to accurately reflect the clinical reality in stroke, patients were not excluded based on stroke type, stroke side, or pre-existing neurological conditions, such as previous stroke. A total of 1159 patients (mean age = 72.5 years (SD= 13.7, range = 18-97), 45.1% female, average education = 11.9 years (SD= 3.82), 7.8% left handed) were included in this study. These participants had an average NIHSS score of 3 (SD=3.82, range =0-22).

Stroke types of the final sample were reported as 909 ischemic, 173 haemorrhagic, 20 TIA, 6 unknown, and 51 not reported. Lesion sides were reported as 466 right, 415 left, 62 bilateral, 148 not visible, and 68 not reported. The average stroke-test interval was 8.1 days (SD=15.4) days. For the multiverse analysis, this participant group was divided into 200 subgroups, each selected based on a plausible combination of lesion location, stroke type, assessment time, and minimum score inclusion criteria (Table 1).

**Table 1:**
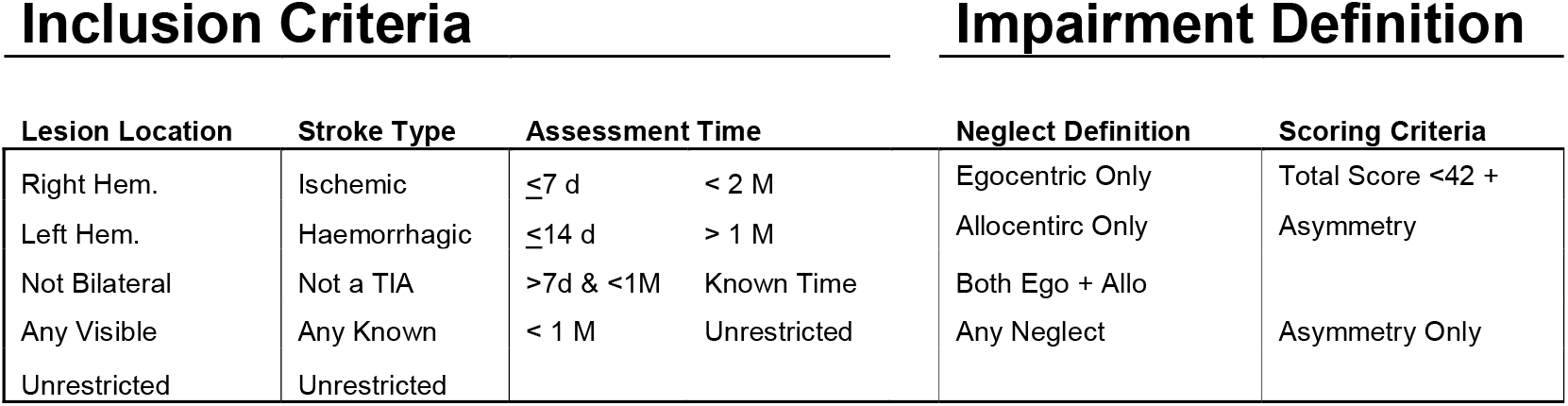
A visualisation of the inclusion criteria and impairment definition factors considered when creating multiverse subsamples with regards to the patient inclusion decisions. For each subsample, one option from each inclusion criteria category was selected, yielding 200 unique patient groups. Each of these 200 groups were then analysed for each of the possible methods for grouping patients with and without significant neglect (impairment definitions). Hem = Hemisphere, Ego = Egocentric, Allo = Allocentric.

## Materials

Each participant in this investigation completed the OCS Cancellation task[23] acutely and at least one standardised recovery outcome measure at 6 months follow-up. Here we considered measures of activities of daily life (including subscales), level of remaining stroke severity, measures of mood, and measures of quality of life. See Table 2 for a full list of all considered outcome measures. The OCS Hearts Cancellation Task has been demonstrated to be highly sensitive to neglect impairment (94.12%, versus the Behavioural Inattention Test Star Cancellation[23]) and to reliably differentiate between allocentric and egocentric visuospatial neglect deficits[22, 24-25].

**Table 2:** A list of the outcome measures considered within this investigation. N represents the number of participants for which data was available for each outcome measure. The Time column reports whether the assessment was delivered during acute hospitalization or at 6-month follow up. SIS = Stroke Impact Scale, NIHSS = National Institute of Health Stroke Scale, ADL = Activities of Daily Living, IADL = Instrumental Activities of Daily Living, HADS = Hospital Anxiety and Depression Scale. Max scores which are marked with a star denote tests in which a higher score represent worse outcome. These scores are inverted in all subsequent analyses. SIS total is the combined total of all SIS subtests. SIS ADL Total is the combined total of both SIS ADL measures.

| Measure | Max Score | Time | N | Mean (SD) |
| --- | --- | --- | --- | --- |
| SIS Total [26] | 300 | Follow-up | 465 | 222.3 (46.3) |
| SIS ADL Total [26] | 90 | Follow-up | 462 | 66.2 (17.5) |
| SIS Physical Weakness [26] | 20 | Follow-up | 451 | 14.5 (4.5) |
| SIS Memory [26] | 35 | Follow-up | 465 | 28.8 (6.2) |
| SIS Mood [26] | 45 | Follow-up | 462 | 32.7 (6.4) |
| SIS Communication [26] | 35 | Follow-up | 460 | 31.0 (5.0) |
| SIS Mobility [26] | 45 | Follow-up | 464 | 34.2 (9.8) |
| SIS ADL[26] | 45 | Follow-up | 462 | 39.2 (10.1) |
| SIS Affected Hand [26] | 25 | Follow-up | 449 | 17.5 (6.9) |
| SIS ADL 2 [26] | 45 | Follow-up | 457 | 27.4 (9.05) |
| NIHSS[27] | 42 | Acute | 319 | 4.3 (3.8) |
| Quality of Life[28] | 20 | Follow-up | 519 | 15.2 (3.1) |
| HADS Depression[29] | 21* | Follow-up | 491 | 5.5 (4.0) |
| HADS Anxiety[29] | 21* | Follow-up | 484 | 6.2 (3.9) |
| Carer Strain Index[30] | 13* | Follow-up | 145 | 7.6 (5.7) |
| Barthel Index[31] | 100 | Acute | 517 | 17.4 (4.1) |

The presence of egocentric neglect severity is scored by subtracting the number of correctly identified targets on the left side of the page from those correctly identified on the right side of the page[23] (Figure One). The cut-offs for egocentric neglect were asymmetry scores of less than −3 or greater than 3[23]. Allocentric neglect impairment on the OCS Cancellation task is scored by subtracting the total number of right-gap false positive responses from the number of left-gap false positives errors made. Cut-offs for impairment are allocentric scores of less than −1 or greater than 1. Some previous studies have employed a more conservative Hearts Cancellation Task Scoring method in which patients are required to have a significant egocentric asymmetry score and a total score of less than 42/50 in order to be classified as having egocentric neglect impairment[9-10]. This investigation employs this alternate scoring method and compares this method to the standard scoring approach (Table 1). For both egocentric and allocentric neglect, negative asymmetry scores denote right-lateralised neglect and positive asymmetry scores denote left-sided neglect.

Given that not all previous investigations have considered both egocentric and allocentric neglect impairment, four different methods for grouping participants into neglect/no neglect groups were employed (Table 1). In the first instance, egocentric impairment alone was considered as the “neglect” category while all other patients were categorised as “no neglect”. Next, allocentric neglect impairment alone was considered. In a third grouping, patients were required to exhibit both allocentric and egocentric neglect to be added to the “neglect” group. In the final instance, patients with any neglect impairment, regardless of the type, were grouped into the “neglect” group.

### Planned Analysis

Multiverse analysis was employed to compare the level of impairment in patients with and without neglect. Multiverse analysis involves multiple permutations of any single base analysis (e.g. regression, t-test, ANOVA). Given this investigation aims to compare a series of continuous variables (e.g. outcome measures) between two categorical groups (neglect vs. no neglect), a t-test base analysis was employed. This base t-test analysis was performed for each possible permutation patient inclusion criteria (n=200), method of neglect impairment definition (n=8), and outcome measure (n=16) yielding 25,600 individual t-tests. Of these tests, only combinations leading to comparisons involving at least 5 neglect and 5 no-neglect participants were considered in subsequent analyses, resulting in a final sample of 18,898 valid comparisons. In line with multiverse methodology, and given that this analysis aims to simulate the findings of all potential individual analyses which could be conducted in isolation, no correction for multiple comparisons are employed[1,8].

First, a specification curve analysis[32] was conducted to evaluate the directionality and robustness of differences between participants with and without neglect. This analysis employed a null curve created by conducting all potential t-test analyses, but with participants randomly allocated to neglect/no neglect comparison groups. The overall generalisability of acute neglect’s relationship with poor recovery outcome was analysed by first evaluating the overall proportion of conducted comparisons met traditional significance thresholds. Subsequent analyses aimed to identify sources of biases within these significant results by comparing average effect sizes (Cohen’s d) across comparisons which employed different selection criteria. All data and code has either been made openly available on the Open Science Framework (https://osf.io/vfnry/) or is available upon request.

## Results

### Specification Curve Analysis

First, a specification curve analysis was conducted (Figure 1). Given the standard alpha level of 0.05, if no effect was present within the analysed data, approximately 5% of comparisons would be expected to yield significant results. However, 33.9% of the conducted comparisons were found to be statistically significant. These significant comparisons had an average effect size of −0.485 (SD=0.327, range=-3.87–0.552, 25^th^ Quantile=-0.523, 75^th^ Quantile=-0.334). This negative overall effect size indicates that as a whole, neglect was found to be associated with poorer performance across the various recoveryoutcome measures.

**Figure 1:**
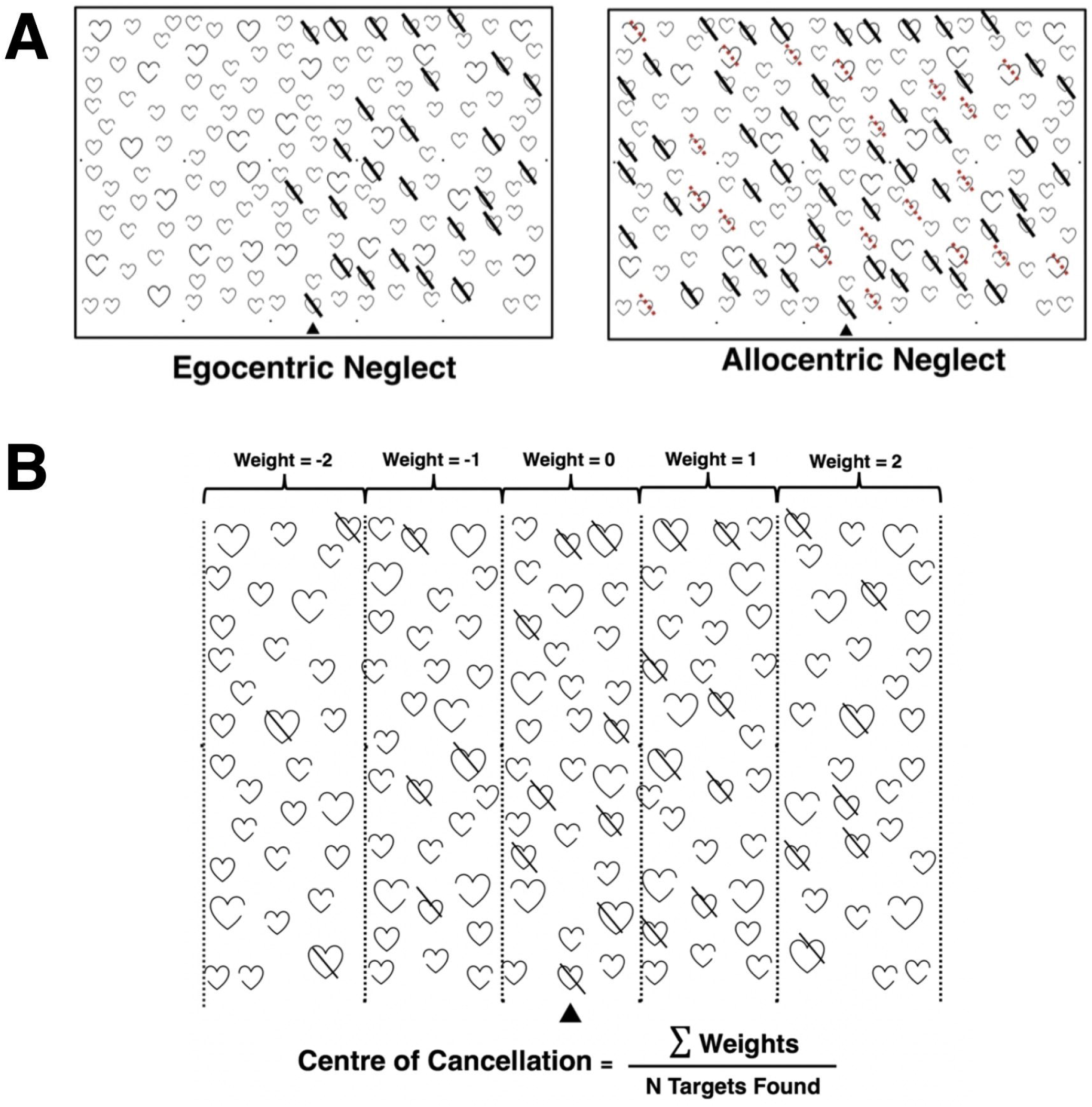
The OCS cancellation task. Panel A presents examples of significant egocentric and allocentric neglect impairment. Panel B presents a visualisation of how the Centre of Cancellation asymmetry metric is calculated.

The robustness of this result was then evaluated by comparing this experimental specification curve to a null curve. The null curve representing randomly-allocated comparisons contained 3272/19200(17.0%) significant results with an average effect size of 0.163(SD=0.387, range =-1.23–1.12,25^th^ Quantile=-0.239,75^th^ Quantile=0.375). 99.9% of statistically significant experimental curve comparisons were found to fall below the null curve 25^th^ quantile boundary (Figure 2). In line with interpretations of the null curve [32], this finding illustrates that the relationship between neglect and poor recovery outcome is highly robust and significant across may possible methodological combinations.

**Figure 2:**
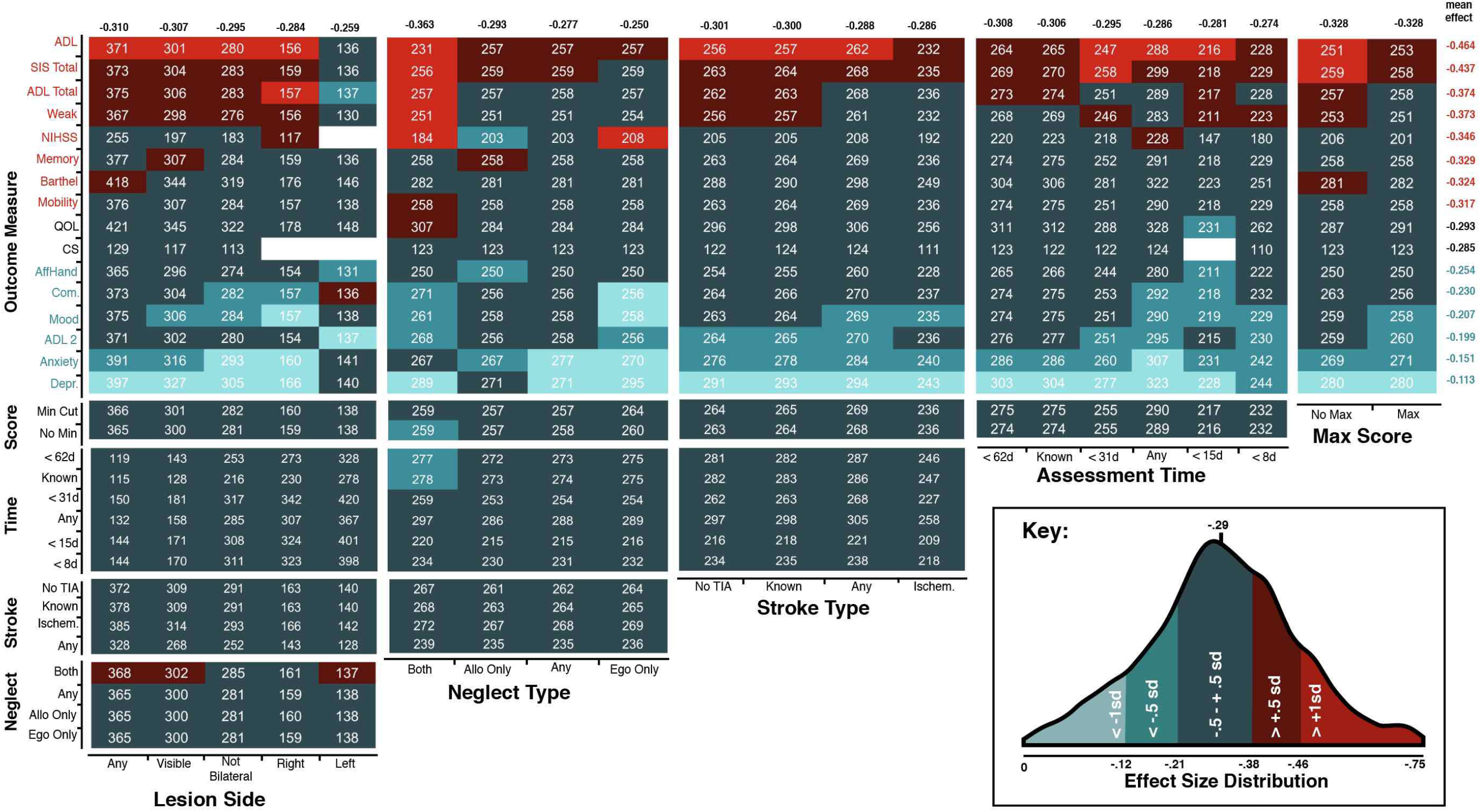
The upper panel presents a specification curve analysis presenting the effect sizes of all experimental and null comparisons ranked in order of size on the x-axis. Significant comparisons are represented in blue, insignificant results are visualised in red, and null curve comparisons are represented in grey. Dotted lines denote the 25^th^ and 75^th^ quantile of the null curve. 99.9% of experimental comparisons fell below the 25^th^ quantile of the null curve, suggesting that neglect illustrates that the relationship between neglect and poor outcome is highly robust and significant across may possible methodological combinations. The lower panels visualise each specific combination of methodological comparisons used in each individual comparison. The average effect size and standard deviation of comparisons using each methodological option are listed to the right of each row. CS = Carer Strain, QOL = Quality of Life, ADL 2 = SIS Instrumental ADL, Com. = SIS communication, AffHand = SIS Affected Hand, Weak = SIS Physical Weakness, Not BL = Not Bilateral, Ego = Egocentric, Allo = Allocentric.

### Evaluating the Impact of Methodological Decisions

Within the robustness of the relationship between neglect and poor recovery outcome, nevertheless significant variance was present within the effect sizes of individual significant comparisons. For this reason, additional analyses were conducted to evaluate the impact of specific methodological choices on resulting effect sizes.

Given that the number of participants with available data within the possible different inclusion criteria choices, neglect grouping factor definitions and across the potential outcome measures was found to vary dramatically, the impact of sample size on the proportion of significant results was explicitly addressed. Comparisons involving less than 100 participants were found to yield only 12.1% significant results while 46.5% of comparisons with 200-299 participants, and 100% of the comparisons including more than 500 participants yielded significant results. Figure 3 presents a visualisation of this known relationship between sample size and proportion of significant results in the present data.

**Figure 3:**
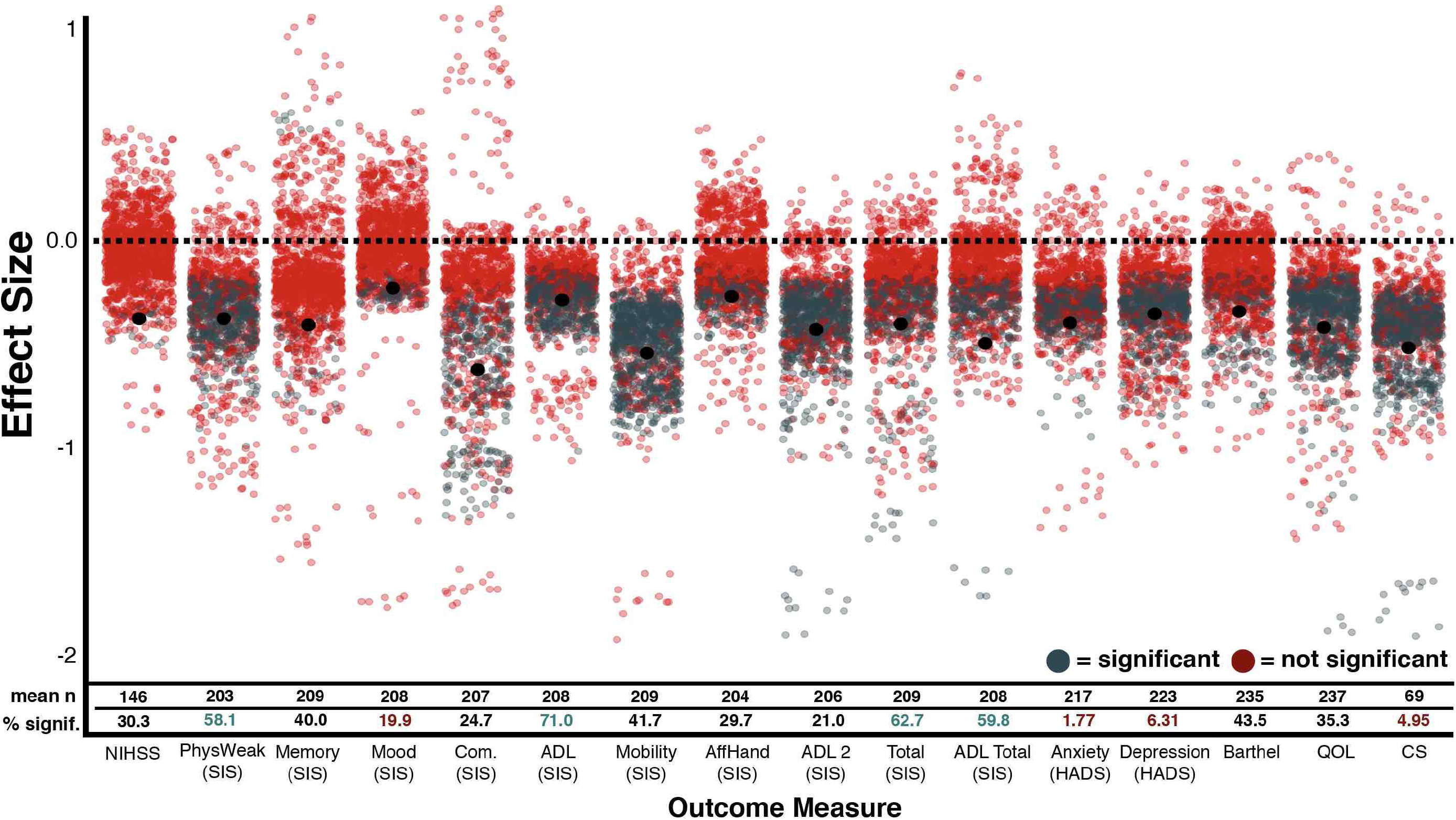
The effect sizes of each conducted comparison grouped by sample size (x-axis). Significant comparisons are presented in blue while insignificant comparisons are in red. Black dots represent the mean effect size of each group. The number of comparisons and overall percent significant results within each group are presented.

Next, the impact of using different outcome measures was evaluated. Average effect sizes were found to vary across outcome measures ranging from −0.28 (HADS Depression) to −0.68 (NIHSS). Overall, comparisons involving HADS Depression (M=-0.28, SD=0.08), SIS IADL (M=-0.32, SD=0.07), SIS Mood (M=-0.40, SD=0.16) and Quality of Life (M =-0.34, SD=0.09) were found to result in the weakest effects and comparisons involving NIHSS (M=-0.68, SD=0.29), SIS ADL (M=-0.63, SD=0.22), SIS Total (M=-0.57, SD=0.41), and SIS Communication (M=-0.55, SD=0.63) were found to be the strongest (Figure 2, Figure 4).

**Figure 4:**
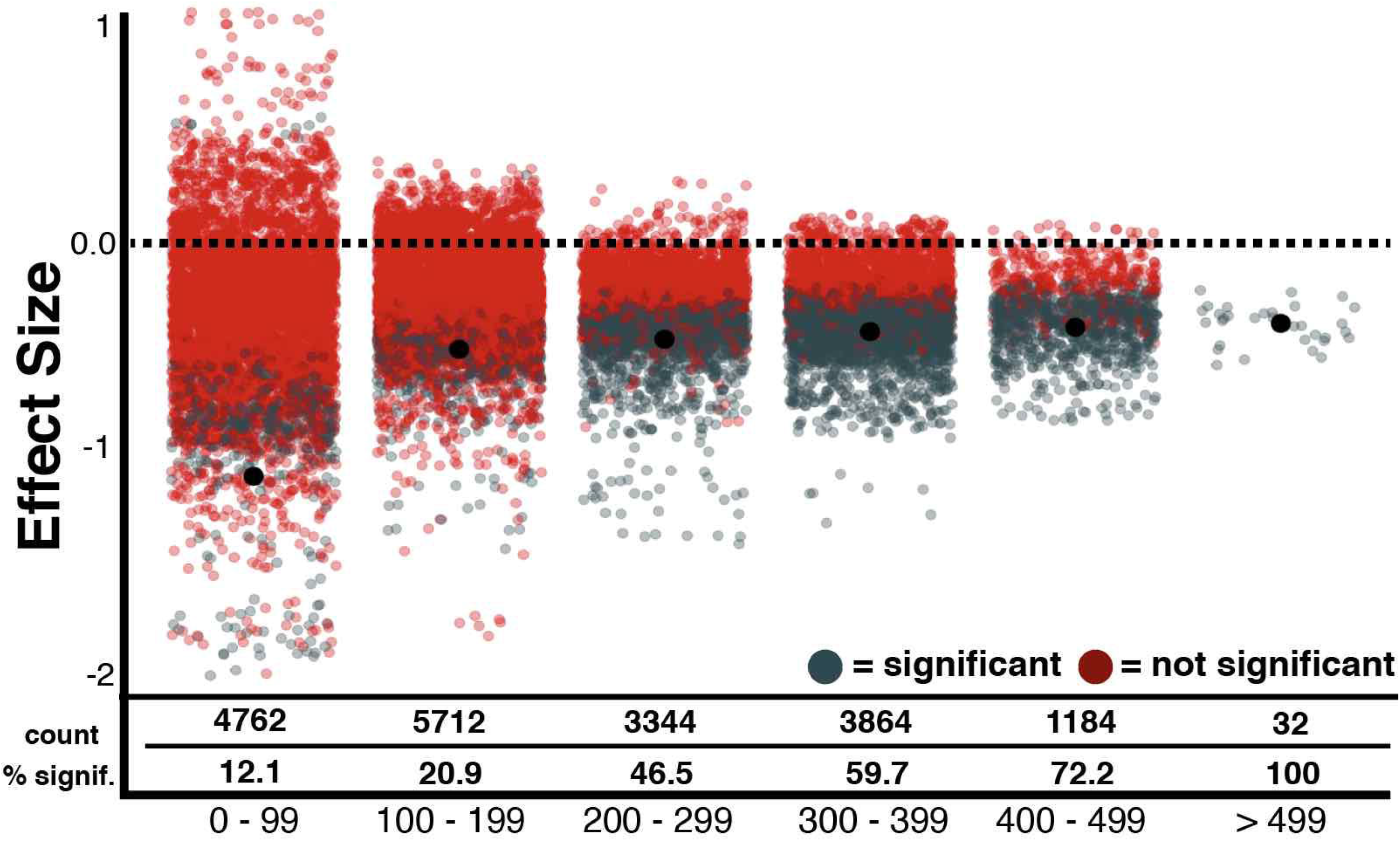
The effect sizes of each conducted comparison grouped by outcome measure (x-axis). Significant comparisons are presented in blue while insignificant comparisons are in red. Black dots represent the mean effect size of each group. The number of comparisons and overall percent significant results within each group are presented. Notably, all outcome measures resulted in an average negative effect size, indicating a robust relationship between the occurrence of acute neglect and poor outcome.

### The Impact of Significant Choice Combinations

Figure 5 presents a visualisation of the impact of specific methodological choice combinations on comparison effect size. Given that many of the conducted comparisons involve small sample sizes which are associated with unreliable effect sizes[33-34], only comparisons involving samples of at least 108 participants were included in this comparison (n=13,736). This inclusion threshold was determined by calculating the minimum sample size needed to detect the average significant effect size (−0.485) with a power of 0.80.

**Figure 5:**
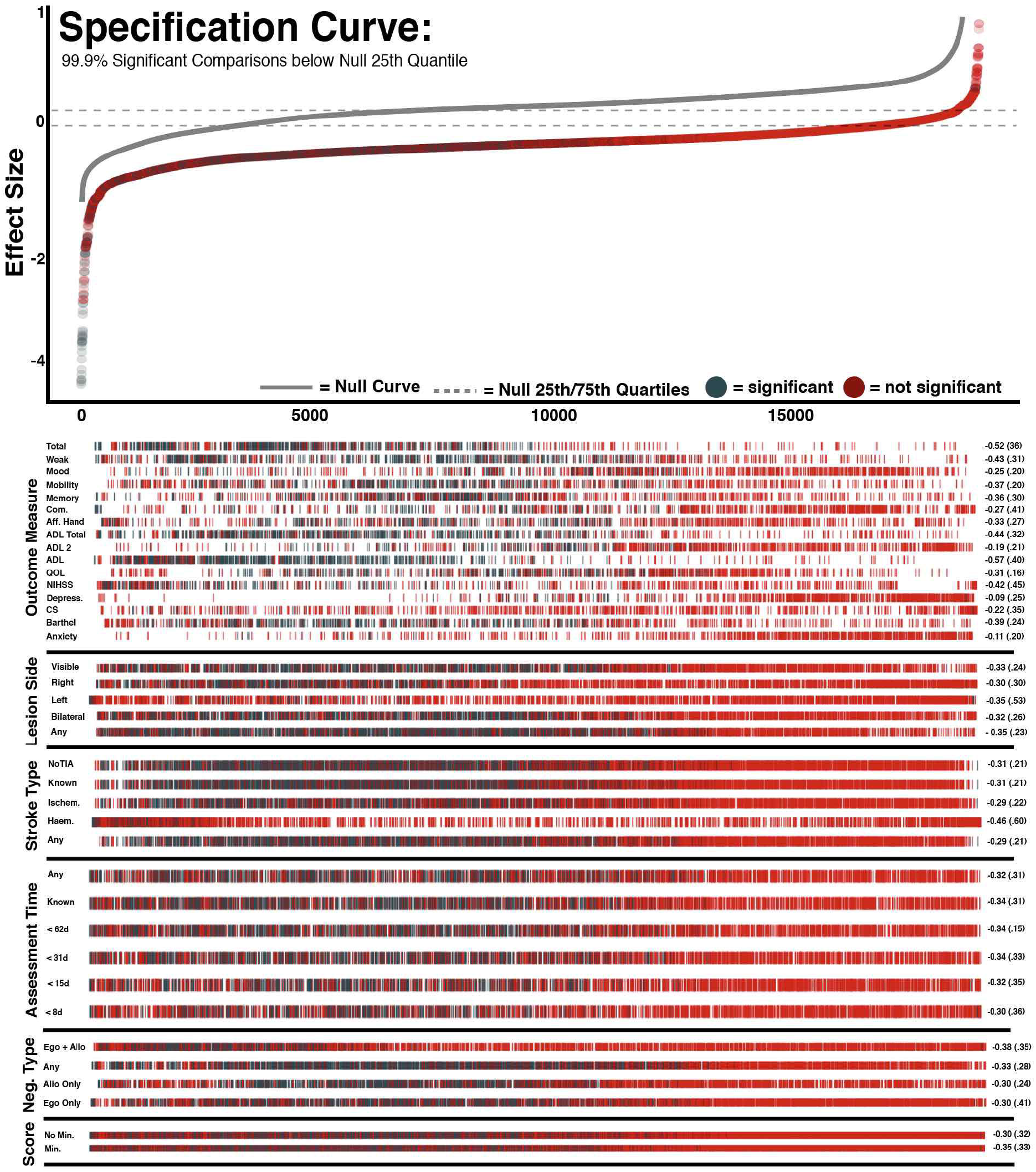
A visualisation of the impact of individual methodological choices on effect size. Each box represents the combination of two specific methodological choices listed on the x and y axes. Box colour represents the average effect size of comparisons which employed each specific combination of choices (See Key). Boxes are labelled with average sample size. The average effect size of each choice across all conducted comparisons is also reported. Red total values represent average effect sizes which are significantly higher than the overall mean effect and blue values represent average effects which are significantly lower than the mean effect as determined by t-tests. Only comparisons which are sufficiently powered to detect the average significant effect size (−0.56) are included in this figure (n > 105).

Overall, the choice of outcome measure was found to have the greatest impact on comparison effect size across choice combinations with the average effect size ranging between −0.464 (SIS ADL) and −.0.113 (HADS Depression). Notably, these consistently negative average effect sizes demonstrate that acute neglect was associated with poor performance across all employed outcome measures. Other selection criteria led to similar ranges in average effect size. Comparisons which included all patients, regardless of lesion type resulted in stronger effects (mean effect=-0.310) than those which included patients with only right (−0.284) or left (−0.259) hemisphere damage (Figure 4). Similarly, analyses which considered allocentric neglect impairment generally resulted in stronger effects than those which considered only egocentric neglect impairment (−0.250). Comparisons which required participants to have significant asymmetry scores as well as total scores of <42 in order to be classified as exhibiting egocentric neglect (mean=-0.328) resulted in stronger average effect sizes than those which only required significant asymmetry scores (mean=-0.268). Assessment time and stroke type and selection criteria resulted in comparatively little variance within average effect sizes.

## Discussion

The broader purpose of this investigation was to demonstrate the utility of multiverse analysis techniques for evaluating the strength and generalisability of effects documented within neurological populations. For this purpose we employed a specific dataset recently used to investigate the relationship between acute visuo-spatial neglect and later functional/recovery outcomes[35]. With the multiverse analysis, we investigated the effect of choice of inclusion criteria, patient group definitions (here presence/absence of acute visuospatial neglect) and choice of outcome measure to represent ‘impact’. The multiverse analysis demonstrated that the predictive relationship between acute visuospatial neglect and poor recovery outcome is a particularly robust effect present across a range of outcome measures, patient groups, and scoring criteria. However, the variations in the strength of this effect across comparisons provides key information on the impact of researcher choices.

This analysis confirms that it is critically important to ensure neurological studies are adequately powered. In this investigation, comparisons employing sample sizes less than 100 were significantly less likely to detect effects which were reliably present when larger cohorts were considered. The risks associated with employing small sample sizes have been well-documented[36-38]. Conducting neurological investigations which are not powered to produce informative null results risks using valuable time and financial resources inefficiently.

Methodological choices relating to outcome measure used, stroke lateralisation, neglect categorisation criteria, and scoring method also created substantial variance within resultant effect sizes. Patients with neglect were found to have comparatively worse outcomes across each of the 16 outcome measures employed within this investigation. However, the average effect size of these comparisons varied dramatically between outcome measures. Neglect was found to have the weakest average impact on mood and the communication subscale of the Stroke Impact Scale. In contrast, the strongest impact was found on measures assessing Activities of Daily Living. Interestingly, employing SIS ADL resulted in the strongest average effect sizes while employing the SIS IADL measure resulted in one of the weakest. The SIS ADL sub-measure aims to investigate mainly self-care actives and includes questions about personal hygiene, household chores, and continence. Conversely, the SIS IADL measure assesses more participation-based metrics and includes questions pertaining to social activities, hobbies, and interpersonal relationships[26]. The results of the present investigation suggests that neglect may impact more heavily on self-care activities than on relational or participation-based activities.

Comparisons which included all patients, regardless of lesion type resulted in stronger effect sizes than those which included patients with only right or left hemisphere damage. Similarly, the precise definition of neglect impairment employed within these comparisons was found to impact resultant effect sizes. Comparisons which exclusively considered egocentric neglect were found to result in weaker effect sizes than comparisons which also considered allocentric neglect. Most interestingly, analyses including only participants exhibiting both egocentric and allocentric neglect resulted in stronger effect sizes than those which included egocentric or allocentric patients alone. These findings align with previous research suggesting that the relationship between acute neglect and poor longer term outcome may not be uniform across all patients with neglect, but may instead vary significantly across different subtypes of neglect[9,24].

The multiverse results ultimately suggest that the strongest relationship between neglect and poor recovery outcome was found when neglect presence/absence was defined on the basis of demonstrating both ego-and allocentric neglect and activities of daily living were chosen as the outcome measure. It is possible this relates to an underlying factor of severity of neglect[10]. Additionally, in order to produce clinically useful information, it is critically important to employ a representative sample of neglect including patients with right and left lesions and egocentric and allocentric impairment. The inclusion of more representative samples will facilitate results which can be generalised to the stroke population as a whole, rather than to only a specific subset of patients.

From this example, we extrapolate that multiverse analysis provides a novel method for evaluating the robustness of effects which could prove particularly useful for neurological studies as it enables researchers to distinguish between large-scale drivers of phenomena and delicate, ungeneralizable effects which are dependent on the exact methodological techniques employed[1], [8], [39]. Multiverse analysis enables researchers to isolate the impact of specific methodological decisions and evaluate the generalisability of their findings to wider patient populations.

Finally, with a growth of open data policies we suggest that multiverse analysis techniques have the potential to inform the design of future confirmatory studies, in an enhanced approach, where in addition to systematic reviews and meta analyses (which present the highest level of evidence[40]), multiverse analysis on existing datasets can present an enhanced opportunity to identify potential sources of bias within existing literature.

## Conclusion

This investigation demonstrates the utility of multiverse analysis techniques within neurological research with an example analysis of post-stroke impact of visuospatial neglect. The multiverse analysis makes explicit effects of researcher choices with respect to patient participant inclusion criteria, diagnostic criteria and primary outcome measure.

## Data Availability

All data and code has been made openly available on the Open Science Framework (https://osf.io/vfnry/).

## Acknowledgements

We would like to express our sincere gratitude and admiration to the late Prof Glyn W Humphreys, who initiated the OCS work and led the OCS-Care study. The OCS study was supported by the National Institute for Health Research Clinical Research Network. We also acknowledge the contributions to data collection and curation for the OCS data made by Ms Ellie Slavkova, Ms Grace Chiu, and Ms Romina Basting.

## Funding

This work was funded by Stroke Association UK awards to ND (TSA2015_LECT02; TSA 2011/02) and MJM (SA PGF 18\100031).

### Abbreviations

OCS: Oxford Cognitive Screen
NIHSS: National Institute of Health Stroke Screen
SIS: Stroke Impact Scale
ADL: Activities of Daily Living
HADS: Hospital Anxiety and Depression Scale

